# Traditional Chinese medicine: A Bayesian network model of public awareness, usage determinants, and perception of scientific support in Austria

**DOI:** 10.1101/2021.12.24.21268331

**Authors:** Michael Eigenschink, Luise Bellach, Sebastian R. Leonard, Tom E. Dablander, Julian Maier, Fabian Dablander, Harald H. Sitte

## Abstract

**Introduction:** Despite the paucity of evidence verifying its efficacy and safety, traditional Chinese medicine (TCM) is expanding in popularity and political support. Decisions to include TCM diagnoses in the International Classification of Diseases 11th Revision (ICD-11) by the World Health Organization (WHO) and campaigns to integrate TCM into national healthcare systems have occurred whilst the public perception and usage of TCM, especially in Europe, remains undetermined. Accordingly, this study investigates the popularity, usage patterns, perception of scientific support for TCM, and its relationship to homeopathy.

**Methods:** A cross-sectional survey was performed in Austria and data from 1382 participants were analysed. A Bayesian network model retrieved partial correlations indicating distinct associations between sociodemographic determinants, complementary and alternative medicine (CAM) usage patterns, readiness to vaccinate, and TCM related variables.

**Results:** TCM was broadly known by the Austrian population (89.9% of women, 90.6% of men), with 58.9% of women and 39.5% of men using TCM between 2016 and 2019. 66.4% of women and 49.7% of men agreed with TCM being supported by science. We found a strong positive relationship between the perceived scientific support for TCM and trust in TCM-certified medical doctors. Moreover, perceived scientific support for TCM was negatively correlated with the proclivity to get vaccinated. Additionally, our Bayesian network model yielded distinct associations between TCM-, homeopathy-, and vaccination-related variables.

**Conclusion:** TCM is widely known within the Austrian general population and actively used by a substantial proportion. However, a crucial disparity exists between the commonly held public perception that TCM is scientific and findings from evidence-based studies. As public opinion towards TCM, and the proclivity to use it, are promoted through institutionalisation and official acknowledgement, it would be critical to sustain and support the distribution of unbiased, science-driven information by governmental institutions and policymakers to encourage informed patient-driven decision-making.

**Strengths and limitations of this study:**

- This is the first study to comprehensively explore the usage patterns and sociodemographic associations of TCM in a European population, not based on data deriving from the seventh round of the European Social Survey.
- We are the first study on CAM usage patterns to graphically explore and report data using a Bayesian Gaussian copula graphical model—thereby, retrieving distinct partial correlations.
- We provide an up-to-date summary of TCM, set forth our findings at a geopolitical scale and highlight that the discrepancy between the paucity of evidence underpinning most TCM modalities and the international promotion of TCM is also reflected in the widely-held public perception that TCM is supported by science.
- Due to the retrospective character of our cross-sectional survey, answers are naturally prone to recall and response bias.
- Our sample is skewed towards the young, people with higher levels of education, and shows a relative underrepresentation of males. Therefore, we post-stratified our sample using representative data from Austria’s federal statistical office “*Statistik Austria*” as a robustness check.

## INTRODUCTION

Traditional Chinese medicine (TCM), an alternative medical practice sculpted by Taoism and Confucianism, was first documented over two millennia ago in the compendium *Huangdi Neijing*.^*1*^ As a core feature of China’s cultural heritage, TCM is predominantly practised in Asia but has gradually been introduced to the international medical community due to China’s opening to the West and its determined promotion by the Chinese Communist Party.^2-4^ The apogee of this expansion can be seen in the World Health Organization’s (WHO’s) decision to include TCM diagnoses in its 11th revision of the *International Classification of Diseases* (ICD-11).^5 6^ This controversial decision follows a lengthy campaign to integrate complementary and alternative medicine (CAM) services into national health care systems through the *WHO Traditional Medicine Strategy 2014–2023*.^7^ Notably, the origin of this document raises concerns over conflicts of interest as its development was supported technically, logistically and financially by the *People’s Republic of China*, the *Government of the Hong Kong Special Administrative Region*, and the *Hong Kong WHO Collaboration Centre for traditional Medicine*.^7^

Despite acquiring international recognition, TCM is founded on a doctrine divergent from a modern understanding of science; invoking the “life-energy” qi^8^, a network of channels through which qi passes called meridians^9^, and the metaphysical forces of yin and yang, which are claimed to be imbued into all entities.^1 10^ In practice, while some randomised controlled trials (RCTs) point towards an effect of TCM therapies on various clinical outcomes^11-14^, meta-analyses and Cochrane-reviews have concluded that the overall data landscape either yields low-quality evidence or is not sufficient to support regular clinical usage.^15-19^ Of note, the largest proportion of TCM-related RCTs examine the efficacy of acupuncture, and effects seem to dissipate upon usage of adequate control group designs.^20-22^ Thus, empirical evidence behind many TCM therapies and diagnostics is deemed poor.^23^

This notion is opposed throughout China where the discrepancy between recognition and evidence is being circumvented through state-run efforts to criminalise criticism of TCM and encourage doctors to prescribe TCM alongside conventional therapeutics.^24^ Whereas abroad, the subsidisation of international TCM education bolsters the promotion of this highly remunerative—$58.9 billion— industry.^24 25^

Compounding the concerns that surround this practice is the endorsement of animal products.^26^ TCM is claimed to treat a plethora of diseases like stroke, myocardial infarction, convulsions, and pain syndromes^27 28^, and an array of animals and their derivatives are used to this end. Pangolin scales, tiger bones, seahorses, rhino horns and bear bile are among a few of the items that have sparked international outcry due to reports of animal rights violations, poaching of endangered species, and the incentivisation of animal trafficking.^28-32^ Indeed, seizure numbers of animal-derived medicinal products totalled 40,434 during 2018, in the EU alone, with Thailand and China constituting the largest exporters.^33^

Today, TCM is exported to over 160 countries^34^ with the US and the EU spending billions of dollars on products.^35 36^ With the establishment of verified TCM schools teaching acupuncture and herbal medicine in North America, Australia and Europe^25^, there are strong reasons to suggest that TCM has spread beyond Asia and established a foothold in these continents.^37^ This growth in popularity sparks concerns regarding patient safety. Contrary to other CAM practices like homeopathy, TCM endorses the application of pharmacologically active substances. However, compared to conventional therapeutics, traditional remedies and phytotherapeutics are not subject to the same regulatory requirements and stringent testing.^38 39^ Due to their non-purified nature and reportedly disparate compositions, concentrations of desired active agents may vary greatly and contamination with potentially poisonous substances might occur.^40-43^ Indeed, unwanted interactions with conventional therapeutics and serious adverse events have been reported.^44-47^

Despite a clear demand for high quality research, there exists a paucity of information on TCM usage patterns in European countries compared to the data available from official Chinese government reports or studies deriving from other Asian countries.^48-50^ Moreover, there is considerable data heterogeneity across European studies evaluating CAM usage patterns.^51^ Few studies have evaluated the general public’s attitude towards CAM^52 53^, even fewer focus on the perception of TCM outside Asia^54-56^, and none on the public’s perception of its scientific support. Thus, following two European social survey data analyses conducted by Kemppainen et al. and Fjær et al.^57 58^, this study aims to elucidate the awareness, acceptance, and usage of TCM in a Central European population, as represented by Austria, whilst putting special emphasis on the public’s perception of TCM’s scientific support. Additionally, we also investigate public trust in TCM practitioners because (I) trust in CAM-practitioners has been shown to be enhanced by institutional guarantees^59^, thus is potentially altered by the perception of scientific support, and (II) trust in practitioners was shown to determine patient satisfaction^60^, thereby possibly influencing usage frequency and treatment expenditures.

Finally, we explore the proclivity of TCM usage when in combination with homeopathy; the most prevalent type of CAM in Austria.^61^ Furthermore, we analyse the relationship between TCM usage and the utilisation of vaccines, as data on CAM-users points towards a reduced readiness to get vaccinated when compared to non-users.^62-65^ Ultimately, using a Bayesian graphical model, we explore the multivariate conditional (in)dependence structure of all variables queried in our survey. Thereby, we retrieve partial correlations that indicate distinct associations allowing us to untangle the complex interactions that underlie CAM-usage.

## METHODS

### Data collection

We performed a questionnaire-based cross-sectional study within the Austrian general population. Our study was divided into two phases: (a) an interview-based street survey and (b) an online survey. The questionnaires were designed using the online survey tool “SoSciSurvey”. The street survey was conducted on the 6^th^ of April 2019 by 19 different interviewers on 12 highly frequented and socio-demographically heterogeneous places throughout Vienna, Austria. Interviewers received detailed instructions ahead of their participation and were in constant communication with the organising team. The online survey was uploaded as an article on the 2^nd^ of March 2019 on the website of the most popular daily free newspaper in Austria (named “*Heute*”, translated “*Today*”). The link was open for participation for a total of six days, spanning from the 2^nd^ of March to the 7^th^ of March.

### Procedures

Individuals were approached indiscriminately by interviewers. After informed consent was obtained and electronically recorded, interviewers asked questions and recorded responses according to a pre-defined answer scheme on a mobile device. Interviewers were instructed to remain impartial and cognisant of their speech and body language to minimise bias introduced by intonation, connotation and gesturing. Responses were recorded anonymously. The median duration of the survey was 189 seconds per participant.

The online survey was identical to the street survey, except for minor modifications that were necessary for the survey to be fully comprehensible in the absence of an interviewer. A CAPTCHA was implemented to distinguish true participants from automated responders. After detailed information about data storage and privacy, consent was obtained through a digital form at the beginning of the survey. As SoSciSurvey does not use IP address filtering, thereby enabling the possibility of repeated participation by the same respondent, participants were directly addressed on the first page of the survey and specifically asked to only participate once. Moreover, we used an average relative completion speed cut-off of 2.0 to filter out meaningless, potentially duplicate responses.^66^ Consistent with the street survey, responses were recorded anonymously. The median duration of the survey was 212 seconds per participant. It is probable that the increased time taken for the online survey compared to the street survey was due to the addition of an introductory page, a CAPTCHA, and a finishing page.

### Questionnaire

Designed by our team to combine classical sociodemographic determinants with indicators of CAM usage, the questionnaire especially focused on the utilisation of TCM. It included a total of 21 items: 6 pertained to sociodemographic factors (gender, age, citizenship, level of education, employment, and income), 8 queried TCM directly (awareness, general usage, usage frequency, perception of scientific support for TCM, trust in TCM-certified medical doctors, and TCM expenses), 4 questions focused on additional CAM usage patterns (homeopathy and vaccination hesitancy) and 3 questions were asked to further characterise our sample (study participation, chronic illness, and money spent on conventional medicine). The questionnaire was pilot tested on a heterogenous group of 20 people of varying age and sociodemographic characteristics in consecutive phases. Inputs were considered and the questionnaire optimised between phases. In some instances, the questionnaire was taken in person with the interviewer present to immediately discuss the respondents’ thoughts (“think aloud”). This allowed for optimisation of question comprehensibility, appropriateness, order, and wording to ensure the validity of questionnaire items. The questionnaire can be obtained from online supplementary file 1 in both German and English.

### Statistical analysis

We used a Bayesian graphical model to explore the multivariate conditional (in)dependence structure of our variables. In a graphical or network model, each variable represents a node and an edge is drawn between two nodes if they are conditionally associated, that is, if they are associated after adjusting for all other variables; the edge weight is given by the strength of this association.^67^ The model allowed us to answer questions such as “how strongly is gender associated with TCM usage frequency after adjusting for all other variables?”. The fact that many conditional associations can be handled within a single model and intuitively visualised made this approach ideal for exploring relationships between our variables.

One of the most widely used graphical models is the Gaussian graphical model, which assumes that the data follow a multivariate Gaussian distribution.^67^ The inverse of the covariance matrix parameterises this model, which then yields a matrix of partial correlations when it is standardised. As many of our variables were categorical (e.g., gender and chronic disease) or ordinal (e.g., education and income), they did not follow a Gaussian distribution. To be able to model the dependencies between our variables without making incorrect assumptions about their univariate marginal distributions, a Gaussian copula was used.^68 69^ The Gaussian copula graphical model^70 71^ allowed us to separate the modelling of the dependency between variables (which is achieved through the Gaussian copula) from modelling the marginal distributions (which we do without assuming a particular parametric form^69^). Similar to the Gaussian graphical model, the Gaussian copula graphical model is parameterised by an inverse covariance matrix which, when standardised, yields a matrix of partial correlations that encode the dependencies between our variables; the key difference is that now the different univariate marginal distributions of our variables are respected. It is this partial correlation matrix, as well as the simple correlation matrix, that we have focused on in our analysis.

Quantifying uncertainty is an important part in any statistical analysis because it guards against overconfidence. Bayesian inference provides a principled and practical approach to quantifying uncertainty, therefore we employed it in our analysis. Bayesian inference requires specifying a prior distribution over parameters, which then gets updated to a posterior distribution when combined with data. We assigned a recently introduced Matrix-F distribution to the inverse covariance matrix in our Gaussian copula graphical model^72 73^. While the Matrix-F distribution constitutes a more flexible prior compared to the widely used Wishart prior, we note that with sample sizes as large as ours, the prior has little influence on the posterior. We used the R package BGGM for our analysis.^73^ We report summaries of the posterior distribution for all partial correlations as well as marginal correlations in the Gaussian copula graphical model. Furthermore, the extent to which the variance of each variable can be explained by all other variables is reported.^74 75^ Note that we did not conduct a statistical power analysis because our goal is not to test hypotheses and control the false positive and false negative error rates. Instead, our goal is to quantify the uncertainty across the estimated (conditional) relationships between variables. Our Bayesian method naturally accounts for uncertainty and guards against overconfidence.

As for data exclusion, 75 participants were excluded from all analyses due to missing and inconclusive data. Moreover, 19 participants were excluded from the analysis depicted in table 1. For TCM-related analyses, 232 individuals who reported unawareness of TCM or indicated inconsistency in their knowledge of TCM, homeopathy, or acupuncture, and 13 participants who had skipped analysis-relevant questions were excluded. Additionally, 34 participants were excluded for not providing information about their medical expenses in our analysis of the personal financial cost of TCM. Moreover, for the Bayesian graphical model, we imputed the income group of 156 participants and the medical expenses of the 34 participants who chose not to provide the respective data using the R package mice.^76^

**Table 1.** Overiew of CAM usage *An overview of male and female CAM usage examining the relative frequencies of five criteria: 1) TCM awareness, 2) usage of TCM, 3) usage of acupuncture, 4) usage of homeopathy, 5) homeopathy propagation*.

| Gender | Knows TCM | Has used TCM | Has used acupuncture | Has used homeopathy | Has recommended homeopathy |
| --- | --- | --- | --- | --- | --- |
| Female | 89.9% | 58.3% | 61.9% | 71.6% | 43.4% |
| Male | 90.6% | 51.2% | 48.6% | 45.1% | 18.7% |

Of note, as our sample was skewed towards the young and more educated population and suffered from a relative underrepresentation of males, we post-stratified it, using data deriving from “*Statistik Austria*”^77^, Austria’s federal statistical office. All results displayed in this paper (except figure 1) are derived from the post-stratified sample. For reasons of transparency, all raw descriptive statistics can be found in the R-script uploaded in our GitHub (*https://github.com/fdabl/TCM-Analysis*) repository. Because a way to use post-stratification in the estimation of the Bayesian Gaussian copula graphical model does not currently exist, we post-stratified manually, subsampling our dataset with replacement using post-stratification weights and estimating the graphical model on the subsampled data. We repeated this procedure 250 times. Combining the estimates from these runs yielded a post-stratified graphical model. The graphical model and figures that are based on the raw data can be found in the supplementary file 2.

**Figure 1.**
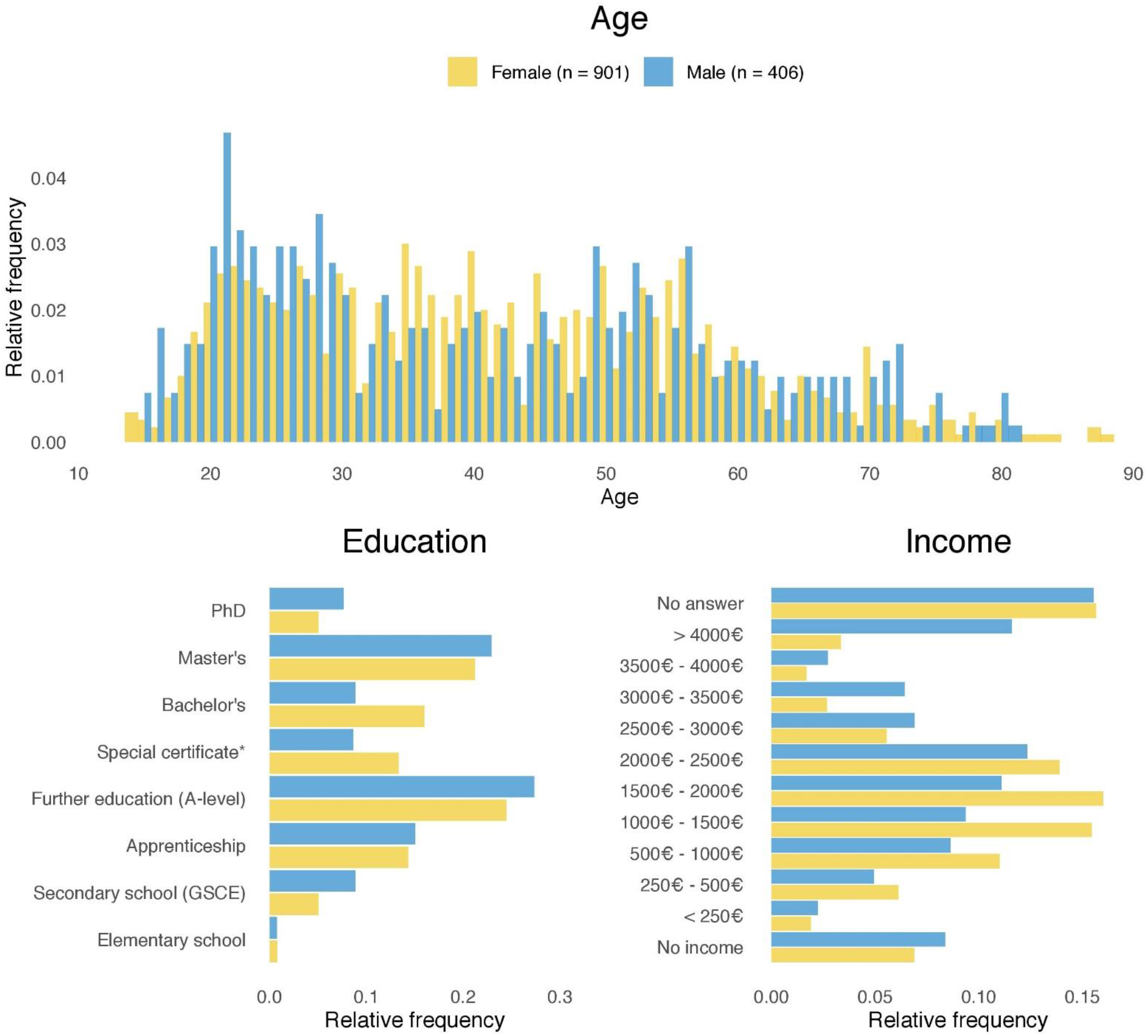
The distribution of (A) age, (B) highest level of education attained, and (C) average monthly income in euro (€), by gender. Blue bars and yellow bars represent male and female participants, respectively. *A-level with additional education in economics, cooking, etc.

All statistical analyses were conducted using the statistical computing language R version 4.0.0.^78^ The R-script can be obtained from the online GitHub repository.

### Patient and public involvement statement

Other than giving feedback on the questionnaire, neither patients nor the general public were actively involved in (I) the design and (II) the conducting of the study as well as (III) the reporting of outcomes and (IV) dissemination of research results.

## RESULTS

A total of 1382 participants completed either the online (*n*=918) or street (*n*=464) survey. After exclusion of missing and inconclusive data^66^, a total of 1307 participants were eligible for data analysis.

### Demographic analysis

Our study population consisted of 901 women and 406 men with a mean age of 41.7 (SD: 15.6) and 40.5 (SD: 16.9) years, respectively. The majority of participants (68% women and 67% men) were employed at the time of data collection (figure 1). 42.2% of women and 39.4% of men had a university degree and 32.1% of women and 47.2% of men reported a monthly income exceeding 2000 euros (excluding 141 women and 63 men who chose not to report their income bracket). Moreover, 45.3% of women and 38.7% of men suffered from a chronic disease. Compared to the Austrian general population, men were strongly underrepresented in our sample population. Additionally, the sample was skewed towards the young with more people in the 15–29 year age bracket (36.7% vs 21.5% for men and 26.8% vs 19.4% for women) and correspondingly less in the 65–84 age bracket (10.6% vs 17.8% for men and 8.6% vs 20.6% for women). The participants also had higher levels of education compared to the Austrian general population: only 8.9% of men and 5.0% of women in our sample reported lower secondary education as their highest level of education, compared to 16.9% and 22.0% in the general population.^77^ Similarly, the proportion of men and women in our sample reporting to have completed a Bachelor’s, Master’s, or PhD qualification is 39.4% and 42.2%, respectively, compared to 12.7% and 12.5% in the general population.^77^

Thus, using data deriving from “*Statistik Austria*”^77^—Austria’s federal statistical office—we post-stratified our sample in order to more closely represent the Austrian general population in terms of gender, age, and education. An analysis of TCM awareness and usage, acupuncture usage as well as homeopathy usage and propagation is presented in table 1.

### TCM-related analysis

Regarding the frequency of TCM usage, 58.9% of women and 39.5% of men used TCM at least once between 2016 and 2019. An examination of participants reporting high usage frequencies revealed a gender imbalance with 31.8% of women compared to 24.9% of men reporting usage at least five times during the same three-year period. 48.6% of women and 30.3% of men used acupuncture at least once in the past three years, with 22.0% of women and 17.8% of men using it at least five times. Further statistics are provided in table 2.

**Table 2.** Analysis of acupuncture and TCM usage between 2016 and 2019 *The number of times male and female participants have used (A) acupuncture or (B) TCM between 2016 and 2019*.

|  | Gender | 0 | 1 – <5 | 5 – <10 | 10 – <20 | ≥20 |
| --- | --- | --- | --- | --- | --- | --- |
| **TCM usage** | Female | 41.1% | 27.1% | 12.1% | 9.3% | 10.4% |
|  | Male | 60.5% | 14.5% | 8.4% | 5.6% | 10.9% |
| **Acupuncture usage** | Female | 51.4% | 26.7% | 8.0% | 7.1% | 6.9% |
|  | Male | 69.7% | 12.4% | 7.4% | 7.3% | 3.1% |

Furthermore, we evaluated the perception of scientific support for TCM and trust in TCM-certified medical doctors. 66.4% of women and 49.7% of men “mostly agreed” or “completely agreed” with TCM being scientific. This trend is congruent with 83.3% of women and 70.2% of men who “mostly agreed” or “completely agreed” with TCM-certified medical doctors being trustworthy alongside 7.2% of women and 18.8% of men who either “did not agree” or “rather disagreed”. Further information regarding the perception of scientific support for TCM and trust in TCM-certified medical doctors can be obtained from table 3.

**Table 3.** Analysis of the perception of scientific support for and trust in TCM-certified medical doctors *Male and female (A) perception of scientific support for TCM and (B) trust in TCM-certified medical doctors. Results conform to a Likert scale in which “do not agree” indicates a perception of weak scientific support and distrust, and “completely agree” indicates perception of strong scientific support and trust*.

|  | Gender | Do not agree | Rather disagree | Partly agree | Mostly agree | Completely agree |
| --- | --- | --- | --- | --- | --- | --- |
| **Perception of scientific support for TCM** | Female | 8.0% | 7.9% | 17.6% | 25.5% | 40.9% |
|  | Male | 18.1% | 10.3% | 21.8% | 26.1% | 23.6% |
| **Trust in TCM-certified medical doctors** | Female | 4.3% | 2.9% | 9.5% | 20.4% | 62.9% |
|  | Male | 12.4% | 6.4% | 11.0% | 21.5% | 48.7% |

In order to better understand the personal financial cost of TCM, we evaluated the most recent three-year history of participant’s medical and TCM expenses. 52.4% of women and 66.9% of men spent less than 100€ on TCM-related therapies, while 14.2% of women and 8.3% of men spent more than 750€. Additional information regarding conventional medicine and TCM expenses can be obtained from table 4.

**Table 4.** Summary of TCM expenses and conventional medical therapy expenses between 2016 and 2019 *The (A) TCM therapy expenditures and (B) total medical expenses of male and female participants between 2016 and 2019. Results are shown in euro*.

|  | Gender | 0€ | 1– <100€ | 100– <250€ | 250– <500€ | 500– <750€ | 750– <1000€ | ≥1000€ |
| --- | --- | --- | --- | --- | --- | --- | --- | --- |
| **TCM expenses** | Female | 42.5% | 9.9% | 12.8% | 13.2% | 7.5% | 5.4% | 8.8% |
|  | Male | 59.9% | 7.0% | 13.9% | 5.3% | 5.6% | 5.1% | 3.2% |
| **Medical expenses** | Female | 6.7% | 13.0% | 13.9% | 15.9% | 15.3% | 8.7% | 26.5% |
|  | Male | 11.4% | 15.7% | 16.7% | 20.2% | 7.9% | 10.3% | 17.8% |

### Bayesian graphical model

A Bayesian Gaussian copula graphical model was used to explore (conditional) associations between the ascertained variables.^69 73^ Figure 2 shows the partial and marginal correlation networks, with edge sizes indicating the posterior mean of the (conditional) associations. The marginal correlation network provides information about how strongly any two variables are related without taking any other variables into account. The partial correlation network, on the other hand, indicates how strongly any two variables are associated if all other variables are taken into account. Naturally, partial correlations are weaker than marginal correlations, as indicated by the reduced edge size observable in figure 2. Below, we focus on partial correlations with posterior means and 95% credible intervals, as shown in figure 3.

**Figure 2.**
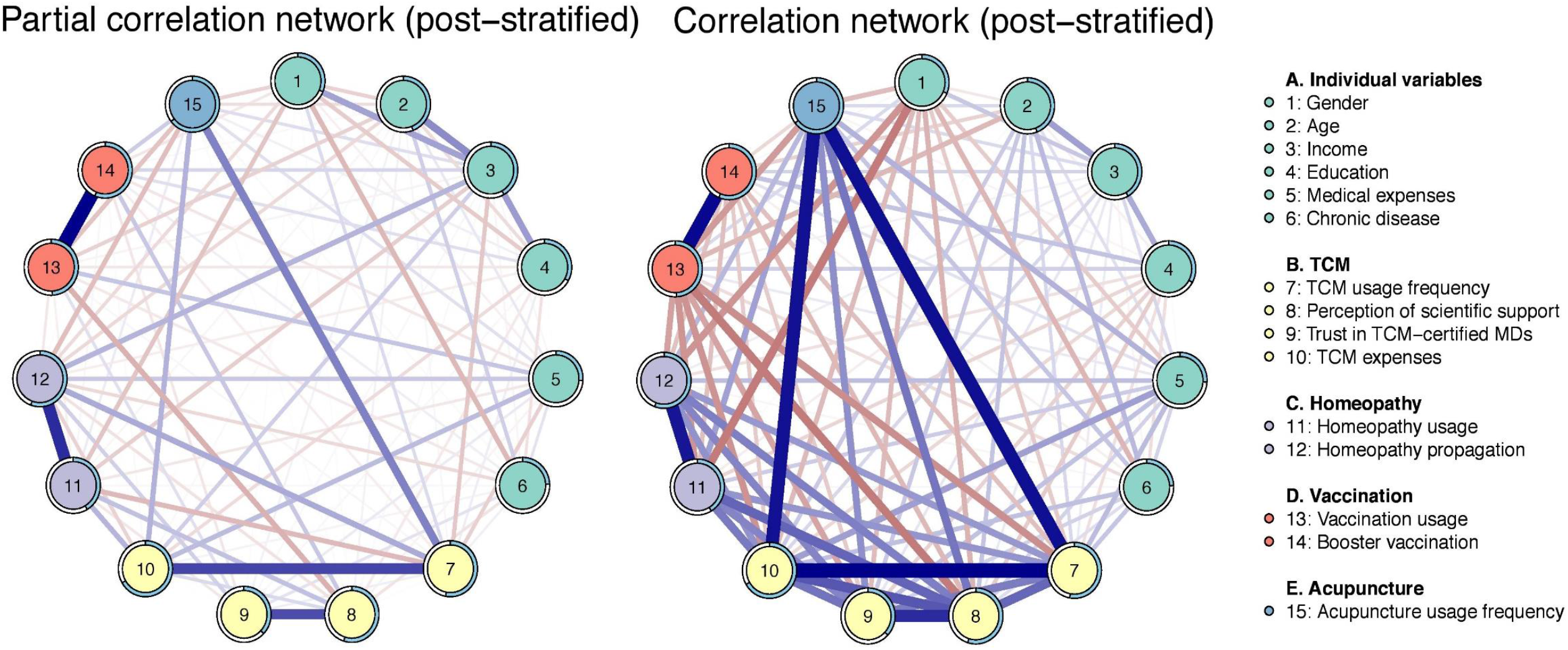
Bayesian network model depicting (A) partial correlations and (B) marginal correlations of variables obtained through the cross-sectional survey. Blue edges indicate positive (partial) correlations, while red edges represent negative (partial) correlations. The circles around the nodes give the posterior mean of the explained variance for the respective variable (more precisely depicted in figure 4).

**Figure 3.**
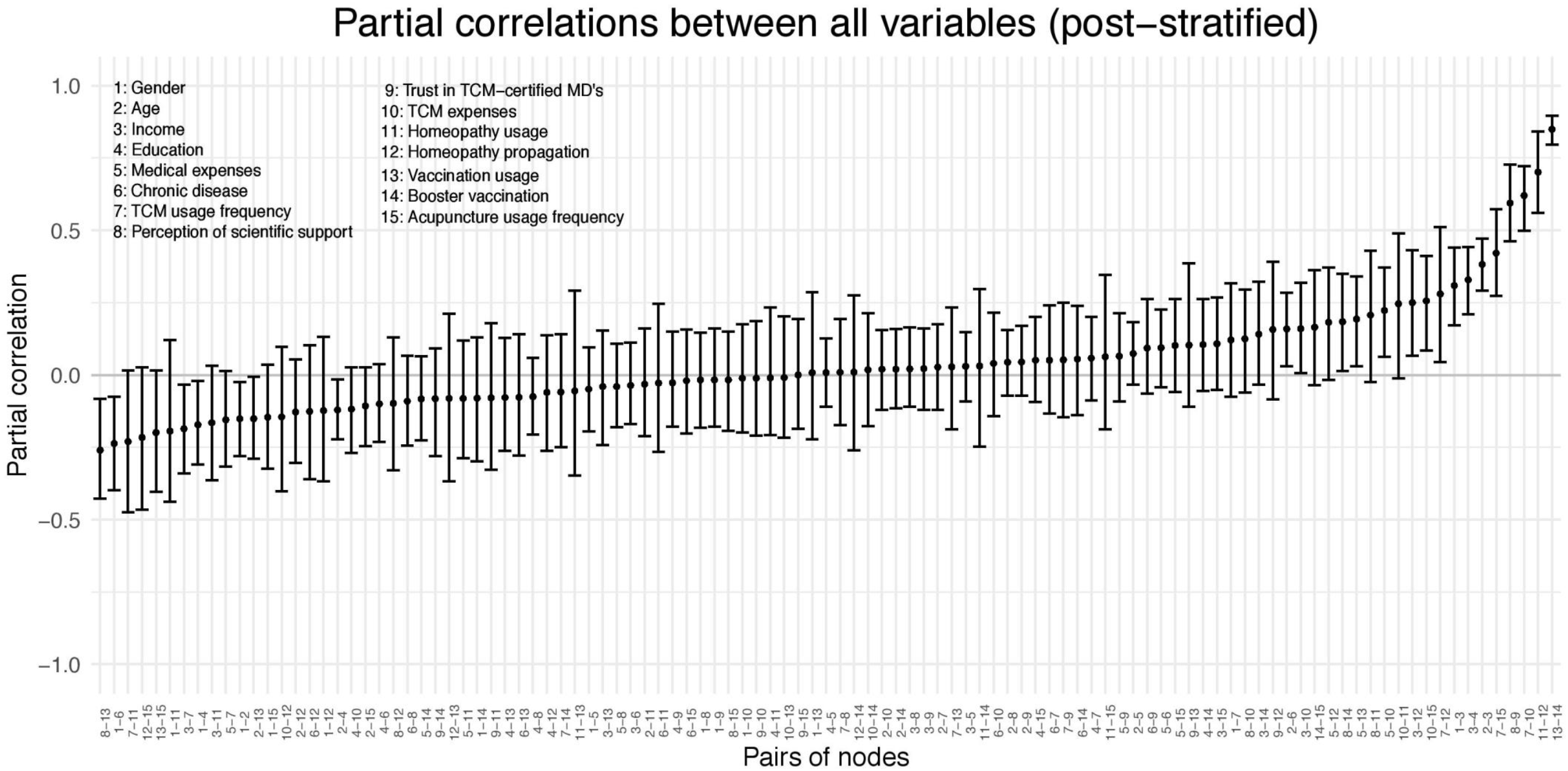
Detailed summary of partial correlations observed in the Bayesian network model for all included nodes. Partial correlations are summarised with posterior means and 95% credible intervals.

Currently, a standardised way does not exist to use post-stratification in the estimation of Bayesian Gaussian copula graphical models. We therefore post-stratified manually, subsampling our dataset with replacement using post-stratification weights and estimating the graphical model on the subsampled data.

Strong positive partial correlations between vaccine usage [13] and utilisation of booster vaccines [14] (ρ = 0.85, 95% CI = [0.80, 0.90]), homeopathy usage [11] and propagation of homeopathy [12] (ρ = 0.70, 95% CI = [0.56, 0.84]), as well as the frequency of TCM usage [7] and money spent on TCM [10] (ρ = 0.62, 95% CI = [0.50, 0.72]) were observed. Interestingly, a strong positive partial correlation was also found between the perceived scientific support for TCM [8] and trust in TCM-certified medical doctors [9] (ρ = 0.59, 95% CI = [0.46, 0.73]).

Regarding money spent on TCM between 2016 and 2019 [10], next to the strong positive partial correlation with TCM usage frequency [7], positive partial correlations with acupuncture usage frequency [15] (ρ = 0.26, 95% CI = [0.08, 0.41]), overall medical expenses [5] (ρ = 0.22, 95% CI = [0.06, 0.37]) and income [3] (ρ = 0.16, 95% CI = [0.01, 0.32]) were observed. TCM usage frequency [7] was also positively associated with the utilisation of acupuncture [15] after adjusting for all other variables (ρ = 0.42, 95% CI = [0.27, 0.57]). Moreover, medical expenses between 2016 and 2019 [5] yielded a positive partial correlation with the proclivity to get vaccinated [13] (ρ = 0.19, 95% CI = [0.03, 0.34]). Interestingly, both income [3] and TCM usage frequency [7] showed positive partial correlations with the propagation of homeopathy [12] (ρ = 0.25, 95% CI = [0.07, 0.43] and ρ = 0.28, 95% CI = [0.04, 0.51]).

The usage of vaccines [13] showed a negative partial correlation with perceived scientific support for TCM [8] (ρ = -0.26, 95% CI = [-0.43, -0.08]). Moreover, age [2] also negatively correlated with the proclivity to get vaccinated [13] (ρ = -0.15, 95% CI = [-0.29, -0.01]). Interestingly, income [3] showed a negative correlation with TCM usage frequency [7] (ρ = -0.19, 95% CI = [-0.34, -0.03]), after adjusting for all other variables. A visual depiction of all partial correlations is shown in figure 3.

While the partial correlations in figure 3 show the (conditional) associations between pairs of variables, figure 4 indicates how much variance of each variable can be explained by all other variables in the model.^75^ This is summarised by posterior distributions of the Bayesian R squared measure for each variable.^74^ A larger R squared for a particular variable implies that the other variables are more relevant in predicting its value.

**Figure 4.**
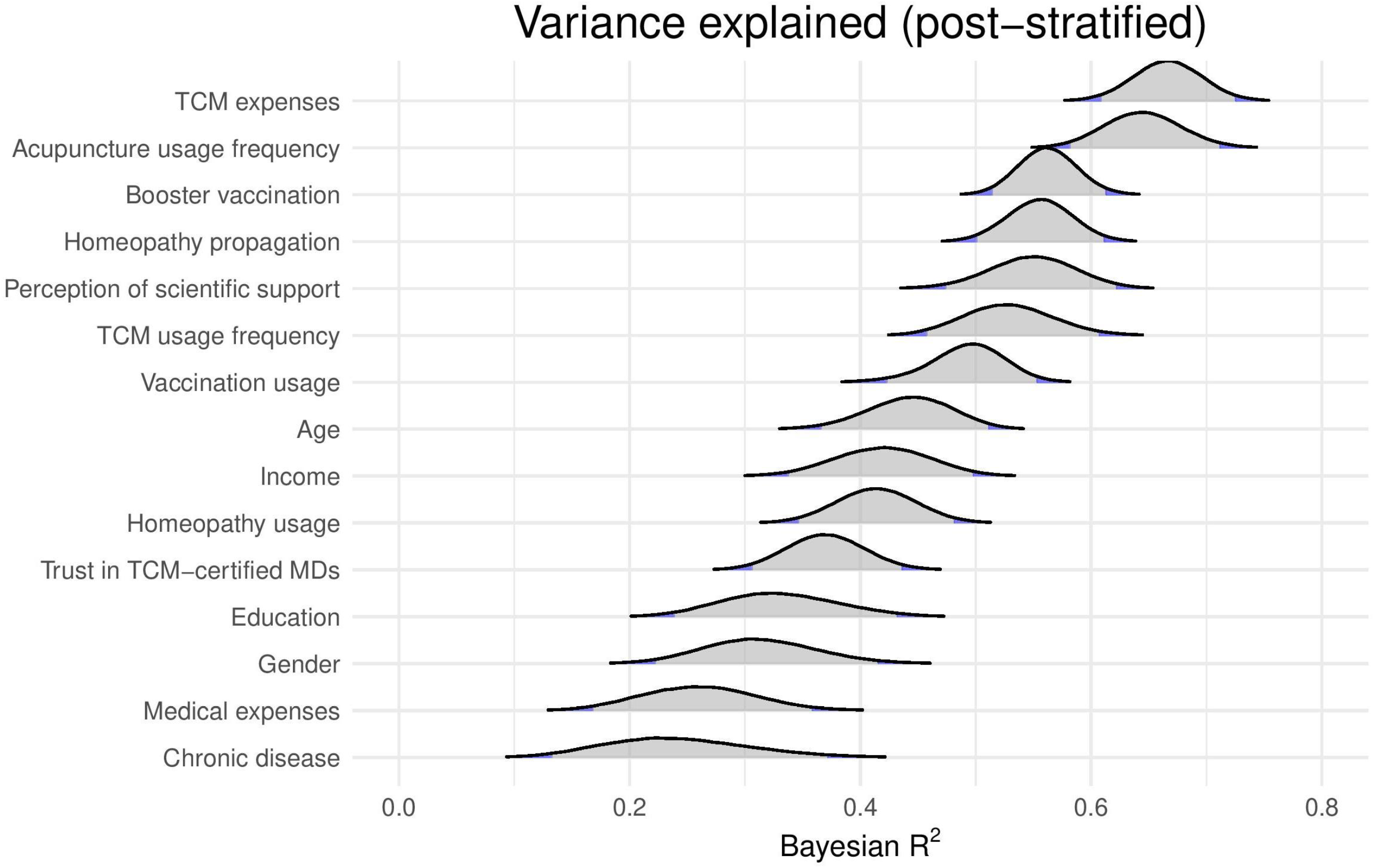
Posterior distributions of the variance explained by all other variables in the model, shown for each queried item.

Notably, TCM expenses and acupuncture usage frequency yielded mean R squared-values of 0.67 (95% CI = [0.61, 0.73]) and 0.65 (95% CI = [0.58, 0.71]), respectively. Moreover, R squared values with a mean greater than 0.50 were observed for the readiness to use booster vaccines (*R*^*2*^ = 0.56, 95% CI = [0.51, 0.61]), propagation of homeopathy (*R*^*2*^ = 0.56, 95% CI = [0.50, 0.61]), TCM usage frequency (*R*^*2*^ = 0.53, 95% CI = [0.46, 0.61]), and perceived scientific support for TCM (*R*^*2*^ = 0.55, 95%CI = [0.47, 0.62]).

Additionally, the usage of vaccines (*R*^*2*^ = 0.49, 95% CI = [0.42, 0.55]), income (*R*^*2*^ = 0.42, 95% CI = [0.34, 0.50]), age (*R*^*2*^ = 0.44, 95% CI = [0.37, 0.51]), and usage of homeopathy (*R*^*2*^ = 0.41, 95% CI = [0.35, 0.48]) presented with mean R squared-values bigger than 0.40. Other sociodemographic determinants yielded lower correlations, e.g., highest level of education (*R*^*2*^ = 0.33, 95% CI = [0.24, 0.43]), gender (*R*^*2*^ = 0.31, 95% CI = [0.22, 0.41]), medical expenses (*R*^*2*^ = 0.26, 95% CI = [0.17, 0.36]), and state of chronic disease (*R*^*2*^ = 0.24, 95% CI = [0.13, 0.37]). Interestingly, while there seems to be a particularly strong partial correlation between perceived scientific support for TCM and the trust in TCM-certified medical doctors, the R squared value of 0.37 (95% CI = [0.31, 0.44]) for TCM trustworthiness is comparatively low—indicating the existence of important additional variables not included in our survey.

## DISCUSSION

The fact that the majority of respondents reported awareness of TCM indicates that the practice has been promulgated throughout the Austrian population. Furthermore, with over a third of men and over half of women reporting to have used TCM between 2016 and 2019, TCM appears to have a notable role as a health care modality within the Austrian population. This is reflected in the public perception that TCM is mostly- or completely supported by scientific evidence, with almost half of men and two thirds of women holding this view. The discrepancy between perception of TCM and empirical support might stem from complex socioscientific interactions and is explored in the paragraphs below.

It must be noted, that even experienced readers of scientific literature might be obfuscated encountering the dichotomy that surrounds TCM. While TCM’s appeals to tradition stand in stark contrast to modern, evidence-based medicine—an observation reaffirmed by empirical evidence— written records of TCM have still led researchers on the path to medical discoveries, namely artemisinin and arsenic trioxide.^23 79-82^ However, it is important to note that these treatments reached clinical acceptance through strict adherence to scientific principles.^79 80 83-85^ TCM’s coalesced scientific existence likely also translates to a clinical and socioscientific setting, further diversifying public perception.

Personal perception of scientific support for TCM was determined by asking the question: “Do you agree with TCM being a science-backed treatment modality?”. While on the surface a simple question, an accurate response depends on a mutual understanding of the definition of science. This raises issues as, even within the domain of science, the word “science” encompasses a number of definitions, including a method of inquiry, a systematised body of knowledge, and a social construct.^86^ In addition, it is plausible that the public interpretation of science extends beyond the formal definitions accounted for in this study. Although there is little evidence available for the Austrian population, this is exemplified in a survey of a British population, in which public attitudes towards science were recorded, and science was found to be collectively understood as a broad and ambiguous topic tantamount to technological advancement. The most popular ideas that emerged when asked to think about science included the three branches of science studied at school, i.e. physics, chemistry, and biology (24%), and simply “*school*” (12%). Other answers included “*chemicals*”, “*white*”, “*coats*”, “*tubes*”, and “*nature*”.^87^ While a survey of this nature calls into question the public’s understanding of the definition of science, it does not report on the level of science literacy. Evidence in this domain suggests that the public have mixed levels of scientific literacy. Indeed, when querying an Italian population, Bucchi and Saracino found that over a third of respondents did not know that the sun is not a planet, electrons are smaller than atoms, or that antibiotics do not kill viruses.^88^

Complicating the issue of ambiguity, the superficial resemblance of TCM and conventional medicine may mislead the public into believing they uphold comparable scientific rigour. This phenomenon has been highlighted by Lobera and Rogero-Garcia, who argue that the scientific appearance of alternative therapies can instil a sense of trust in the layperson.^89^ Overlaying this affect is authority bias, in which experts are credited with competence by virtue of their position. Commonly experienced in a medical setting, it is conceivable that TCM transposes an illusion of medical authenticity through authority bias.^90^ Altogether, TCM’s historical ties in the development of conventional treatments in combination with an imprecise public perception of science; the superficial resemblance of TCM and conventional medicine; and the eminence-based veil through which TCM is communicated^23^ could be contributing factors towards a belief that TCM is sufficiently supported by scientific evidence. Future studies may benefit from pre-defining the term “*science*” with key-words or including qualifying questions to assess science literacy in their survey.

Notwithstanding the disparity between perceived scientific support and the absence of credible evidence that applies to many TCM treatments, the Austrian general public’s trust in TCM-certified medical doctors is high. Our data shows that 83.3% of women and 70.2% of men “mostly-” or “completely trust” physicians with a TCM degree. While efforts have been made to introduce different item scales to standardise the measurement of trust between patients and physicians, high quality data is scarce. Nevertheless, it seems generally accepted to distinguish between two modalities: social trust and interpersonal trust.^91 92^ Covering social trust, a Dutch study conducted by van der Schnee et al. showed trust in CAM to be associated with positive media coverage, recommendation by acquaintances, and personal experience with CAM. Moreover, the analysis showed trust in CAM practitioners to increase with awarded institutional guarantees. 86.3% of participants reported an increase of trust should the CAM practitioners also hold a degree in conventional medicine—a finding corroborated by a study analysing a German population.^59 93^

Analogous to the aforementioned impact of aesthetics on perceived scientific support for TCM, the showcasing of CAM certificates and degrees, as well as partnerships with official CAM associations, appears to enhance trust in TCM practitioners.^59^ This hypothesis aligns with our results since a strong positive partial correlation between trust in TCM doctors and perceived scientific support for TCM was observed—supporting the notion that the perception of TCM’s scientific fundament by the general public might also depend on the institutionalisation, official recognition, and certification of TCM education. Indeed, official TCM degrees can be acquired for substantial amounts of money at multiple universities, TCM schools, and educational institutions in Austria.^94-98^ Moreover, the influence of the scientific community and scientific journals on the portrayal of TCM has to be critically examined. For example, bought special issues in Nature (2011) and Science (2014) lend credibility and visibility to TCM practices.^99 100^

Regarding TCM expenses, our data show that approximately 10% of participants spent more than a total of 750€ on TCM between 2016 and 2019. This finding is interesting since out-of-pocket payments for medical care approximate a total of 901,25€ per capita per anno in Austria.^57 101^ Importantly, Austria employs a compulsory health care insurance system with the possibility of optional private health care. In this context it has to be emphasised that in Austria the coverage of CAM by tax-financed health care insurance has been a topic of political discourse.^102^ Yet, the idea of integrating CAM therapies within national health care services when there is insufficient empirical evidence to validate their efficacy and safety is not unique to Austria. This development is connected to the political and economic dimensions of TCM.^5^ It is a declared goal of the Chinese Communist Party that TCM should be (i) universally accessible for the Chinese population, (ii) attract health tourists to China, and (iii) spread its influence across the world.^2 4 5^ Indeed, the WHO Traditional Medicine Strategy 2014–2023, influenced by the former WHO Director-General Margaret Chan (2006–2017), considers “promoting universal health coverage by integrating T&CM [traditional medicine and complementary medicine] services and self-health care into national health systems”^7^ one of its three main objectives. An inclusion of TCM diagnostic criteria in the ICD-11 thus makes the diagnoses billable to insurance companies, furthering the expansion of TCM as a worldwide business model, while the scientific basis of many TCM treatments is refuted.^5^

## CONCLUSION

An indisputable disparity exists between the public notion that TCM is supported by scientific evidence and the empirical evidence itself, which portrays a widely unsubstantiated alternative medical practice. Arguably, this discrepancy is also mirrored in political decisions that enhance the credibility of CAM by encouraging its institutionalisation and integration within national health care systems. Considering the lack of evidence proving TCM’s efficacy and safety, its endorsement of pharmacologically active agents—and simplified authorisation thereof—as well as the efforts made to criminalise TCM criticism in China, authorities should focus on a critical examination of TCM. Accordingly, treatments supported by empirical evidence should be pursued, and critical discourse based on scientific principles should be fostered. Furthermore, authorities should adhere to strict intellectual honesty, and place an emphasis on improving the scientific literacy of the general public to safeguard informed patient-driven decision-making, especially in the face of potential mis- and disinformation.

### Limitations

As the cross-sectional survey included retrospective components, answers are naturally prone to response- and recall bias. Moreover, as we did not record (I) the number of individuals approached in the street survey and (II) the number of individuals that accessed our online survey, we are unable to provide a survey refusal and response rate. Nonetheless, it must be noted that upon usage of multivariate models, non-response bias does not necessarily alter results.^103^ In order to obtain a larger sample size, an online survey was performed in addition to the street survey. Because the survey was uploaded on the website of Austria’s most popular free newspaper (“*Heute*”), the collected sample might partially reflect the characteristics of its regular readership rather than the general population. Additionally, a relative underrepresentation of male participants can be found in our study population. Thus, we post-stratified our sample with data deriving from *“Statistik Austria”*—Austria’s federal statistical office—in order to more closely represent the Austrian general population. As our data are limited to Austria, future studies are warranted to investigate our findings on an international level.

## Acknowledgements

The authors of the manuscript are particularly grateful for the assistance of the interviewers who voluntarily participated in the creation of our dataset and, therefore, laid the foundation for the study and the present manuscript. Moreover, the authors thank Lisa Krammer and Lukas Dearing for their valuable comments on the questionnaire, and Donald Willliams for answering questions about his BGGM package.

## Author contributions

Guarantor: M.E.; conceptualisation, M.E., L.B., T.E.D., J.M., H.H.S.; methodology, M.E. and T.E.D.; survey, M.E., L.B., T.E.D., J.M., H.H.S.; data analysis, F.D. and M.E.; data curation, F.D. and M.E.; writing—original draft preparation, M.E., L.B., F.D. and S.L.; writing—review and editing, J.M., S.L., F.D. and H.H.S.; project supervision, H.H.S. All authors have read and agreed to the submitted version of the manuscript.

## Patient consent for publication

Not required.

## Ethics statement

After correspondence with the chair of the ethics committee of the Medical University of Vienna, due to the study design, approval by an ethics committee was not required.

## Transparency statement

The lead author affirms that this manuscript is an honest, accurate, and transparent account of the study being reported; that no important aspects of the study have been omitted; and that any discrepancies from the study as planned (and, if relevant, registered) have been explained.

## Data availability statement

The datasets used and/or analysed during the current study are readily available from our GitHub repository: *https://github.com/fdabl/TCM-Analysis*.

