## Supplementary material for "Traditional Chinese medicine: A Bayesian network model of public awareness, usage determinants, and perception of scientific support in Austria": Online supplementary file 1

### Survey evaluating the status and perception of traditional Chinese medicine in the Austrian population

(German introduction and data protection agreement not translated)

1. CAPTCHA:
Please solve the following calculation to prove you are not a robot.
Eight plus three equals (free form)

2. Are you familiar with the term traditional Chinese medicine (TCM)?
Yes
No

3. Have you ever used Traditional Chinese medicine (TCM) or been treated with TCM?
Yes
No
I am unfamiliar with TCM
I do not know

4. How often have you used TCM-related services within the past three years?
0 times
1– <5 times
5 – <10 times
10 – <20 times
20 or more times
I am unfamiliar with TCM
I do not know

5. Do you agree with TCM being a scientifically supported treatment modality?
I completely agree
I mostly agree
I partly agree
I rather disagree
I do not agree
I am unfamiliar with TCM
I do not know

6. Would you trust TCM-certified medical doctors?
I completely agree
I mostly agree
I partly agree
I rather disagree
I do not agree
I am unfamiliar with TCM
I do not know

**7. How much money have you spent on medical treatments (consultations, medication etc.) in the past three years?**
0 €
1 – <100 €
100 – <250 €
250 – <500 €
500 – <750 €
750 – <1000 €
1000 € or more
I do not know

**8. How much money have you spent on TCM (TCM consultations, teas, acupuncture, etc.) in the past three years?**
0 €
1 – <100 €
100 – <250 €
250 – <500 €
500 – <750 €
750 – <1000 €
1000 € or more
I am unfamiliar with TCM
I do not know

**9. Have you used homeopathy in the past three years?**
Yes
No
I am not familiar with homeopathy
I do not know

**10. Have you recommended homeopathy to anyone within the past three years?**
Yes
No
I am not familiar with homeopathy
I do not know

**11. Would you vaccinate yourself or your children?**
Yes
Rather yes
Partly
Rather no
No

**12. Are you conscious about receiving your booster vaccines?**
Yes
Rather yes
Partially
Rather no
No

13. Have you ever used acupuncture?
Yes
No
I am not familiar with acupuncture
I do not know

14. How often have you used acupuncture within the past three years?
0 times
1 – <5 times
5 – <10 times
10 – <20 times
20 or more times
I am not familiar with acupuncture
I do not know

15. How have you come across this survey?
Heute.at
Facebook
Directly sent to me (E-Mail, SMS, etc.)
I participated in a street survey
Other

16. With which gender do you identify?
Female
Male
Other

17. How old are you (in years)?
free form

18. What is your citizenship?
Austria
Germany
Hungary
Romania
Serbia
Bosnia
Turkey
Croatia
Other (free form)

19. What is your highest attained level of education?
Elementary school
Secondary school (GSCE)
Apprenticeship
Further education (A-levels)
Special certificate (A-levels with an additional education in economics, cooking, etc.)
Bachelor´s

Master‘s
PhD

20. Are you employed at the moment?
Yes, I am employed
No, I am unemployed
No, I am retired
No, I am a housewife/househusband
No, I am currently in training
None of the above-mentioned

21. What is your net monthly income? (Monthly income is defined as the amount of money obtained after deduction of taxes. Additionally, please include official sponsorships and other financial support)
I do not have an income
Less than 250 €
250€ – <500€
500€ – <1000€
1000€ – <1500€
1500€ – <2000€
2000€ – <2500€
2500€ – <3000€
3000€ – <3500€
3500€ – <4000€
4000€ or more
I do not wish to comment

22. Are you currently suffering from a chronic disease or have you suffered from a chronic disease in the past three years? (A chronic disease is defined as a disease that develops slowly and lasts longer than four weeks)
Yes
No
