## Supplementary material for "Traditional Chinese medicine: A Bayesian network model of public awareness, usage determinants, and perception of scientific support in Austria": Online supplementary file 2

### Supplementary figures presenting the raw data, that has not been post-stratified (Figure 2-4)

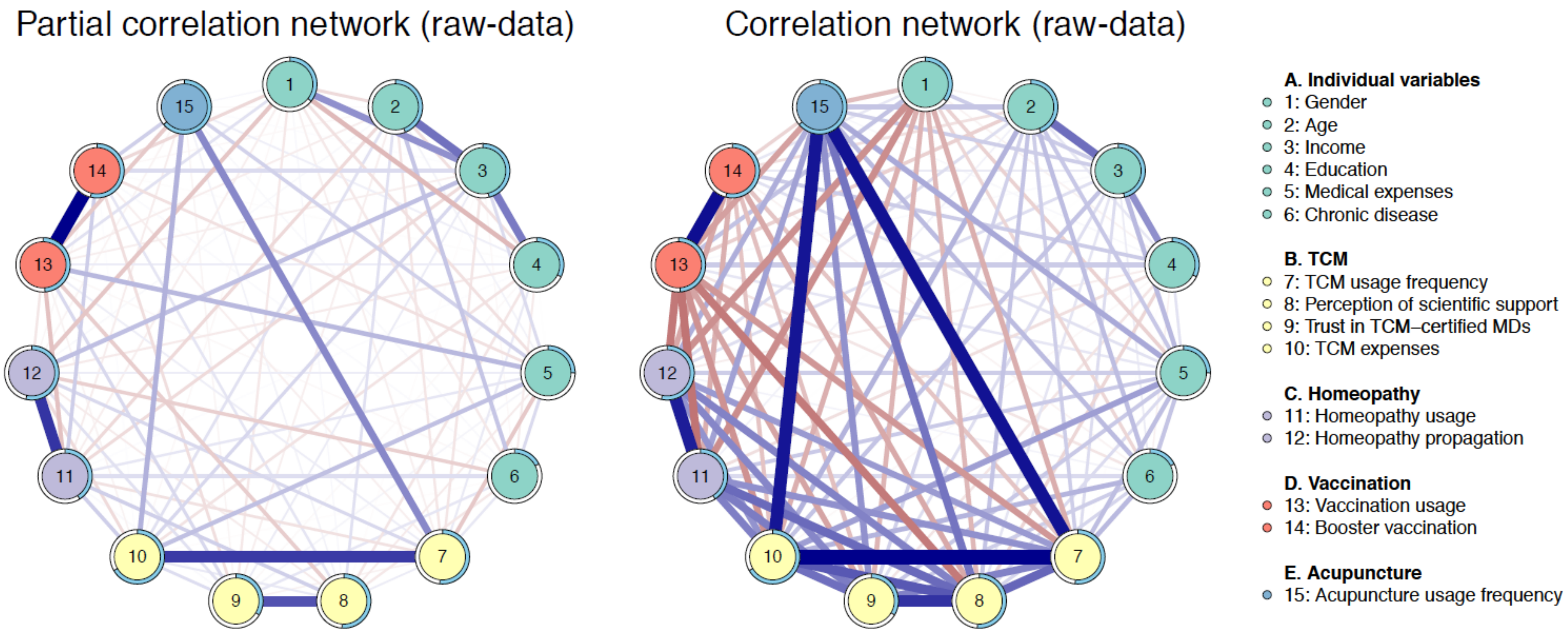


**Figure 2S:** *Bayesian network model of the raw-data depicting (A) partial correlations and (B) marginal correlations of variables obtained through the cross-sectional survey. Blue edges indicate positive (partial) correlations, while red edges represent negative (partial) correlations. The circles around the nodes give the posterior mean of the explained variance for the respective variable (more precisely depicted in figure 4).*

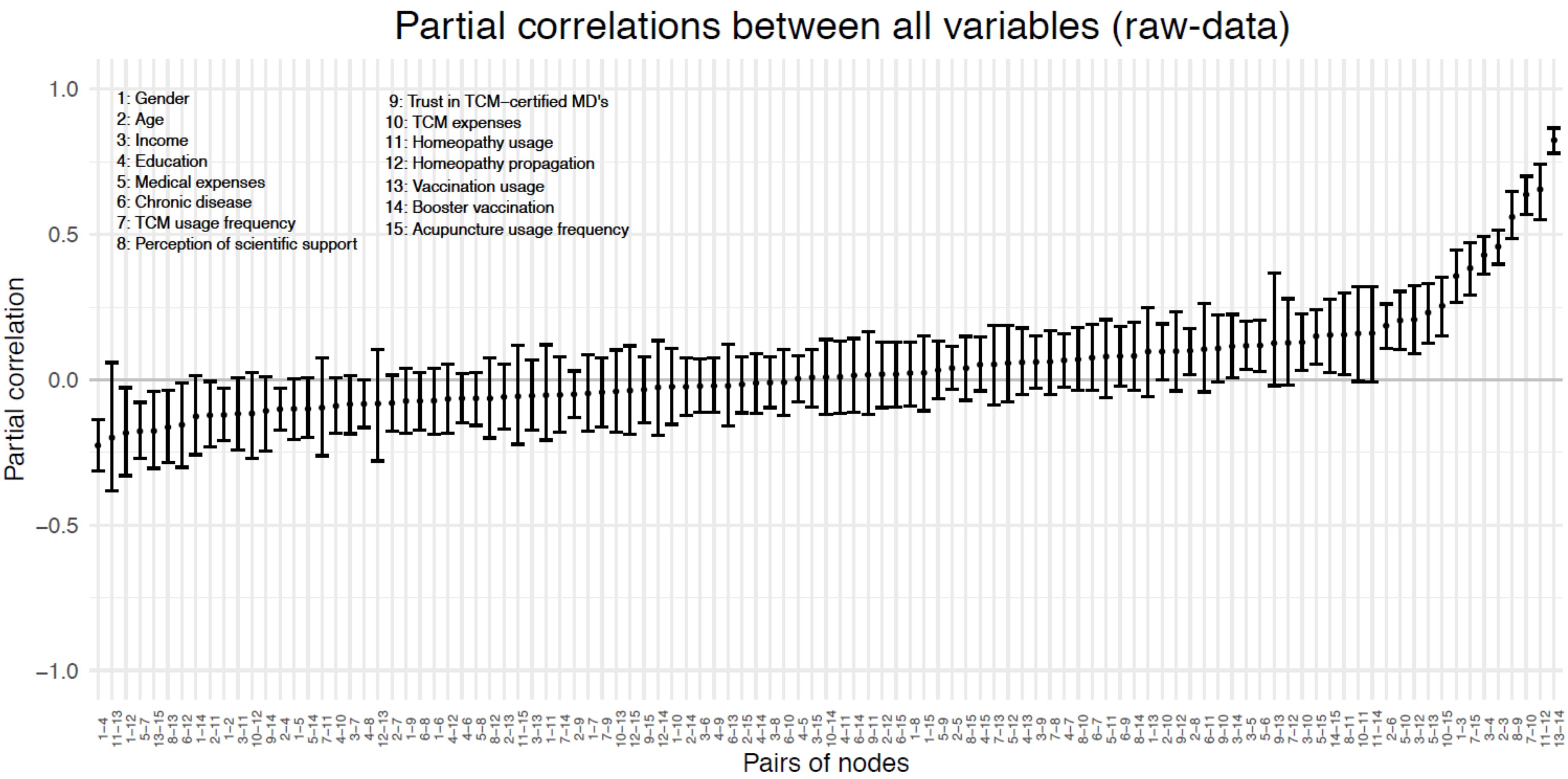


**Figure 3S:** *Detailed summary of partial correlations observed in the Bayesian network model of the raw-data for all included nodes. Partial correlations are summarised with posterior means and 95% credible intervals.*

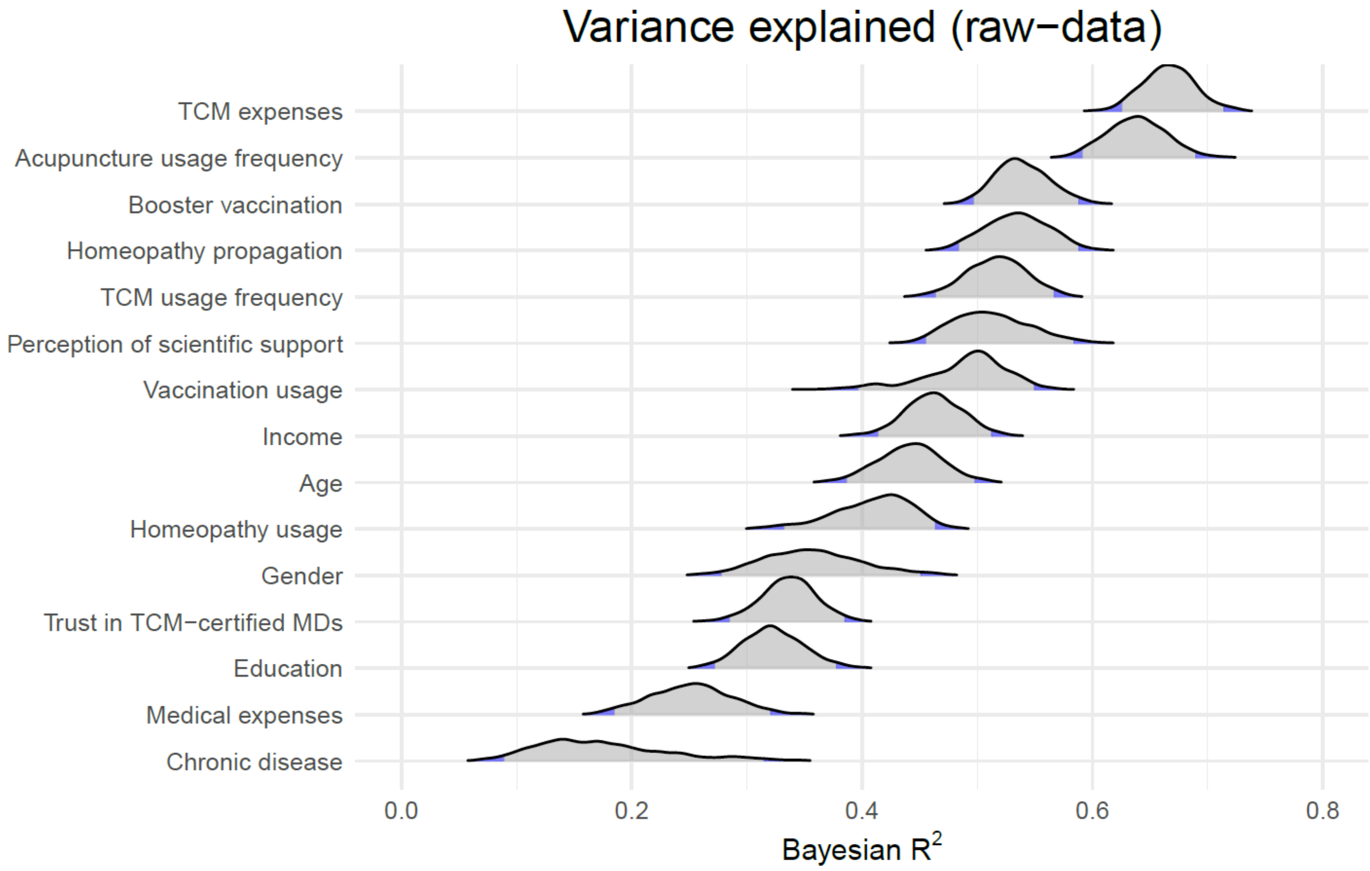


**Figure 4S:** *Posterior distributions of the variance explained by all other variables in the model of the raw data, shown for each queried item.*
